# Effects of vaccination against COVID-19 on the emotional health of Peruvian older adults

**DOI:** 10.1101/2022.01.24.22269781

**Authors:** Christoper A. Alarcon-Ruiz, Zoila Romero-Albino, Percy Soto-Becerra, Jeff Huarcaya-Victoria, Fernando M. Runzer-Colmenares, Elisa Romani-Huacani, David Villarreal-Zegarra, Jorge L. Maguiña, Moises Apolaya-Segura, Sofía Cuba-Fuentes

## Abstract

**Background:** COVID-19 vaccination may reduce anxiety and depression. However, the pandemic significantly impacted the elderly from low-middle-income countries. Therefore, we aimed to estimate the effect of vaccination against COVID-19 on the emotional health of older adults.

**Methods:** We selected a nationally stratified sample of non-hospitalized adults aged 60 to 79 years who intended to receive the COVID-19 vaccine or had already received it during recruitment. We assess the fear, anxiety, and worry about COVID-19, general anxiety, and depression at baseline and after a month. We estimated the adjusted odds ratios (aOR) and 95% confidence intervals (95% CI) for each altered emotional health outcomes in those who had one and two doses, compared with those who were not vaccinated using multilevel logistic regression with mixed effects.

**Results:** We recruited 861 older adults. Loss to follow-up was 20.8%. At baseline, 43.9% had only one dose of the vaccine, and 49.1% had two doses. In the analysis during follow-up, those who had two doses had less fear (ORa: 0.19; CI95%: 0.07 to 0.51) and anxiety to COVID-19 (ORa: 0.45; CI95%: 0.22 to 0.89), compared to non-vaccinated. We observed no effects in those with only one dose.

**Limitations:** Inability to obtain the planned sample size for primary analysis. There is a selection bias during recruitment and a measurement bias because of self-reported vaccination.

**Conclusions:** COVID-19 vaccination with two doses in elders improves the perception of COVID-19 infection consequences. This information could be integrated into the vaccination campaign as its beneficial effect.

**Highlights:**

- Up to 90% of elders in a Peruvian sample had at least one dose of COVID-19 vaccine
- Two doses of COVID-19 vaccine reduced the levels of fear and anxiety for COVID-19
- Only one dose of vaccine didn’t had effect in any emotional mental outcome

## 1. Introduction

The COVID-19 pandemic is having a significant impact on the mental health of people worldwide (Santomauro et al., 2021). Approximately 15% of older adults had a mental disorder before the pandemic (World Health Organization, 2017). But, they have reported greater declines in social communication, exercise, and finances during the last two years than young adults (García-Fernández et al., 2020; Turna et al., 2021). In addition, the high contagiousness of COVID-19 and higher risk of death and complications in the elderly population (Biswas et al., 2021; Tiruneh et al., 2021), may have caused a worsening of sleep quality, well-being, depressive, and anxious symptoms, since the beginning of the pandemic (De Pue et al., 2021; Gerlach et al., 2021). It is estimated that the damage has been especially profound in older adults in developing countries compared to those in developed countries (Babulal et al., 2021). Furthermore, most of the documents of the ministries of health from Latin American countries have not prioritized the strategies or policies that deal with emotional and mental problems during the pandemic, which could cause their impact on these aspects to be greater (Bonilla-Cruz et al., 2020). For example, Peru is one of the countries with the highest mortality rate from COVID-19 per million inhabitants in the world (Taylor, 2021) and has had a significant economic and social impact (Varona and Gonzales, 2021). In this context, high levels of worry, anxiety, and fear of COVID-19 have been described in people living in Peru (Caycho-Rodríguez et al., 2021c; Mejia et al., 2020; Porter et al., 2021).

Vaccination against COVID-19 has meant a change in the pandemic dynamics (Carbone et al., 2021) due to its proven effectiveness in reducing severe cases and deaths from COVID-19 in the general population and older adults (Bernal et al., 2021; Ranzani et al., 2021). Mental health status during the pandemic could be related to COVID-19 vaccination in different ways (Figure 1). First, mood disorders such as stress, depression, and loneliness can decrease the immune system response caused by the COVID-19 vaccine (Madison et al., 2021). In addition, in older adults, having a mental disorder may be associated favorably (Lawrence et al., 2020; Marrie et al., 2021) or negatively (Andrew et al., 2004; Bazargan et al., 2020) with the willingness to be vaccinated against COVID-19. On the other hand, doubts about the COVID-19 vaccine correlate with high levels of anxiety, depression, and post-traumatic stress (Palgi et al., 2021), which can cause acute episodes of anxiety immediately after receiving the COVID-19 vaccine (Hause et al., 2021). Finally, studies in the general population of the United States (Chen et al., 2022; Nguyen, 2021; Perez-Arce et al., 2021) and in health professionals in Turkey (Sugihara et al., 2021) suggest that receiving the COVID-19 vaccine may have a direct effect in reducing levels of anxiety and depression.

**Figure 1.**
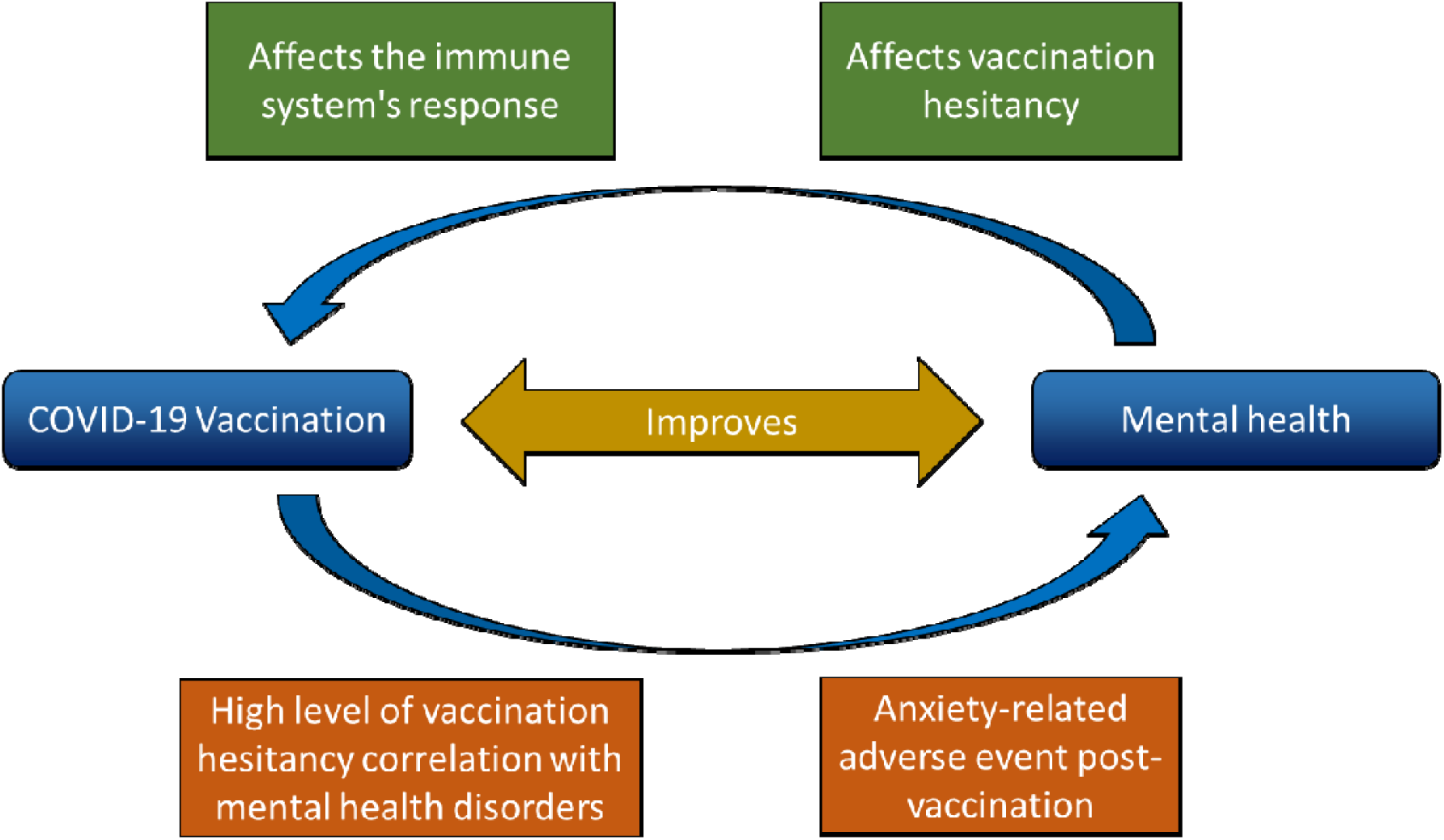
Relationship between vaccination against COVID-19 and mental health in the general population.

Older adults, who are a priority group to receive the COVID-19 vaccine (Brenner, 2021), could also positively affect their emotional health (Kearney et al., 2021; Sapra, 2021). However, this population is underrepresented in studies evaluating this association. A better understanding of the relationship between vaccination against COVID-19 and emotional health would improve knowledge of the positive determinants of health in the population (VanderWeele et al., 2020). The positive effect that vaccination against COVID-19 could generate in the older adult population would imply an improvement in their quality of life during the pandemic, which is how the policies to promote vaccination could be oriented in this direction. So, it is necessary to know how receiving the COVID-19 vaccine could affect the perception of this disease in older adults. The objective of the present study was to assess the effect of receiving the COVID-19 vaccine on emotional health in a representative group of older adults in Peru during the year 2021.

## 2. Methods

### Context

Peru is a country with a fragmented and heterogeneous health system. Among them, the Social Health Insurance of Peru (EsSalud) is one of the most important health systems in the country and is managed by the Ministry of Labor and Employment Promotion (Alcalde-Rabanal et al., 2011). EsSalud gives medical attention to formal workers, retirees, and their families. The executive function of EsSalud is divided through the 29 healthcare networks, representative of each region in the country. EsSalud provides health coverage to almost a third of Peruvians, including more than 1 890 000 older adults, of which 100 000 of them are users of the Centers for the Elderly (CAM, in Spanish) present in each healthcare network in the country (Tenorio-Mucha et al., 2021). The CAMs provide health services that seek to improve the functional, mental, and social capacity of people aged 60 or older affiliated with EsSalud (Romero-Albino and Ortigueria-Sánchez, 2021).

The vaccination process against COVID-19 in Peru, organized by the Ministry of Health, was carried out by age groups, and began on April 16, 2021, with adults over 80 years old. Vaccination started on April 28, 2021, for adults over 70 years old, and on May 27, 2021, for adults over 60 years old. During that period, the BNT162b2 (BioNTech, Pfizer), ChAdOx1-S (Oxford, AstraZeneca), and BBIBP-CorV (Sinopharm) vaccines were available in Peru. However, the Peruvian Ministry of Health indicated that vaccination should be prioritized with BNT162b2 (BioNTech, Pfizer) for older adults with 60 years or older (Ministry of Health, 2021). As result, as of December 29, 2021, 80.8% of older adults who received at least one dose of the COVID-19 vaccine received BNT162b2 (BioNTech, Pfizer), while 10.5% and 8.7% had received the ChAdOx1-S vaccine (Oxford, AstraZeneca) and BBIBP-CorV (Sinopharm), respectively (Ministry of Health, 2021). However, the vaccination of older adults occurred during a political and health scandal in the country, where the inoculation of the BBIBP-CorV (Sinopharm) vaccine candidate under investigation to 470 people was exposed outside the clinical trial in Peru, including health personnel and politicians (Chauvin, 2021; Mayta-Tristán and Aparco, 2021). For several months, this caused a credibility crisis for COVID-19 vaccines, particularly BBIBP-CorV (Sinopharm) (Arango Olarte et al., 2021).

### Study design

Prospective cohort study that aimed to estimate the effect of the COVID-19 vaccine on the following emotional health outcomes of older adults affiliated to EsSalud CAMs: (a) Perception of fear, (b) anxiety, and (c) worry for COVID-19, (d) general depression, and (e) general anxiety.

### Population

We identified non-hospitalized adults aged 60 to 79 years affiliated to EsSalud and registered in the CAM available database at the national level. The database represented 0.5% of all older adults affiliated to EsSalud. We included those older adults who had already been vaccinated or who had planned to be vaccinated against COVID-19 according to the Peruvian Ministry of Health vaccination schedule at a vaccination site in Peru. We excluded adults aged 80 years or older because more than one month had elapsed since the start of vaccination in this age group by the initiation of the recruitment. We also excluded those who had some impediment to adequate communication with the interviewer via the telephone call, were diagnosed with COVID-19 in the last three months, had symptoms related to COVID-19, or refused to participate during the interview. The recruitment period was from May 27 to June 30, 2021.

### Sample size calculation

Based on Cohen et al. recommendation (Cohen, 2013; Lakens, 2013), we consider an effect size of 0.20 standard deviations for the smallest effect (Cohen, 2013) estimated between any of the outcomes (perception score for fear, anxiety, and worry about COVID-19, general depression, and anxiety) and the exposure factor (unvaccinated vs. vaccinated with only one dose or vaccinated with two doses) in older adults. Assuming significance level of 5%, statistical power of 80%, and equal variances between groups, we calculated a minimum sample of 788 older adults. Then, we corrected this value by a factor of 1.2, following the methodology proposed by Vititingghoff E. et al. (Vittinghoff et al., 2012), considering the primary analysis with adjustment for potential confounding variables. Thus, we obtained a minimum sample size of 946 older adults. Finally, we considered a rejection rate of 10% and a loss to follow-up rate of 10%; then, we purposed to invite 1168 participants to the study.

### Sampling

After excluding those who did not meet the selection criteria or did not have identification or contact data, we took 7 685 older adults from the registered CAMs into the database as a sample frame. From them, we chose a randomized and stratified sample for each of the 30 healthcare networks (n=1 686). In addition, the sampling was carried out independently for two age subpopulations: 60 to 69 years (n=846) and 70 to 79 (n=840) years. For each subpopulation, we chose half of the calculated total sample size (allocation ratio 1:1). We decided to choose two age subpopulations because the recruitment period was during the start of vaccination of adults older than 60 to 69 years and one month after the vaccination initiation to adults older than 70 to 79 years (Ministry of Health, 2021). To reduce nonresponse bias, we adjusted sample weights to account for nonresponse using weighting class adjustment (Valliant et al., 2018; Valliant and Dever, 2018).

### Outcomes

We assessed five outcomes in emotional health: Fear of COVID-19, anxiety for COVID-19, worry for COVID-19, general anxiety, and general depression, perceived by the respondents during the last two weeks before to the survey response. Fear of COVID-19 was measured with the Spanish version of the Fear of COVID-19 Scale (FCV-19S) with seven items that are answered on a Likert scale from 1 (Strongly disagree) to 5 (Strongly agree) (Huarcaya-Victoria et al., 2020). This scale measures the emotional and somatic fear response to COVID-19. It has adequate internal consistency (Cronbach’s α = 0.88) and convergent validity with other mental health covariates in two non-probabilistic samples of adults and older adults from Lima, Peru (Caycho-Rodríguez et al., 2021b; Huarcaya-Victoria et al., 2020). The anxiety for COVID-19 was measured using the Spanish version of the Coronavirus Anxiety Scale (CAS), which measures persistent and excessive concern about COVID-19 that is accompanied by physical symptoms (Silva et al., 2020) and has five items that are answered on a Likert scale from 0 (Not at all) to 4 (Almost every day) (Caycho-Rodríguez et al., 2020). It is a unidimensional scale with adequate internal consistency (Cronbach’s α = 0.89 and 0.91), convergent validity with anxiety, and adjustment rates between females and males, and between older adults aged 60 to 65 years and 66 to 86 years. However, estimations in samples of adults and older adults from Lima, Peru, using Item Response Theory models, suggest that the instrument is more reliable in those with high anxiety levels to COVID-19 (Caycho-Rodríguez et al., 2021d, 2021e). The outcome worry for COVID-19 was measured with the scale to measure worry for contagion of the COVID-19 (PRE-COVID-19, in Spanish), which contains six items that are answered on a Likert scale of 1 (Never or rarely) to 4 (Almost all the time). This scale measures the degree of worry for possible COVID-19 infection in the respondent and how this concern affects their state of mind and their ability to carry out their daily activities. It was developed in a non-probabilistic sample of adults between 18 and 50 years of age from Lima and Callao, Peru, and it proved to be unidimensional, to have adequate content validity, internal consistency (ω coefficient = 0.90), and convergent validity with other mental health covariates (Caycho-Rodríguez et al., 2021c). A higher total score on each scale means a higher respondent’s outcome level. Then, we dichotomized the total scores for each outcome and considered the upper quartile as a high outcome level.

The general anxiety outcome was measured with the Spanish version of the Generalized Anxiety Disorder (GAD-2), which contains two items that are answered on a Likert scale from 0 (never) to 3 (almost every day) (García-Campayo et al., 2012). A total score of two or more points can diagnose clinically relevant anxiety in older adults, with 67% sensitivity and 90% specificity (Wild et al., 2014). The general depression outcome was measured with the Spanish version of the Patient Health Questionnaire (PHQ-2), which contains two items that are answered on a Likert scale from 0 (No day) to 3 (Almost every day) (Caneo et al., 2020). A total score of three or more points can diagnose clinically relevant depression in older adults without cognitive impairment with 79% sensitivity and 82% specificity (Boyle et al., 2011).

### Exposure variable

We asked about self-reported COVID-19 vaccination status (no dose, only first dose, and two doses of vaccine) and the time in days since they received each dose of the vaccine.

### Covariables

In addition, we asked about sociodemographic variables, previous mental health diagnosis and treatment, and personal and family history of COVID-19. In addition, we assessed the comorbidity with the Geriatric Comorbidity Index. This index measures the severity degree of 15 clinical conditions, classifying them from 0 to 4 each (0: no disease, 1: asymptomatic disease, 2: asymptomatic disease with treatment, 3: uncontrolled disease despite treatment, and 4: very serious or life-threatening disease). After presenting the severity degree classification and giving simple and standardized examples about each clinical condition, the interviewers asked the responders to identify their current situation for each clinical condition. According to these scores, the degree of comorbidity was grouped into classes: Without comorbidity (all conditions absent), class I (one or more conditions with a severity degree of 1 or less), class II (one or more conditions with a severity degree of 2), class III (one condition with a severity degree of 3), and class IV (two or more conditions with a severity degree of 3, or at least one condition with a severity degree of 4) (Rozzini et al., 2002).

### Data collection

We conducted a pilot with trained interviewers for data collection for one day using a random sample from the sampling frame of approximately 120 participants. During the pilot, we evaluated the data collection capacity of the interviewers and the availability to participate of the selected older adults. Then, we identify and correct deficiencies for formal data collection.

After sampling, we assign an identification code to each selected older adult to facilitate recognition and monitoring within the program. Then, the interviewers contacted the selected participants through telephone calls, using the telephone numbers registered in the CAM database. Previously trained interviewers made the calls and collected data. In case of not answering two calls on two different days, the older adult was excluded from the study. During the call, the interviewers identified the older adult by asking them for their identity document number. Then, the older adult was invited to participate in the study by requesting their verbal informed consent. If the older adult accepted participating in the survey, the interviewer asked for selection criteria. Then, the interviewers collected the baseline data. The data collection interview took approximately 20 minutes.

Then, the follow-up calls were made between July 1st and July 27th, 2021, considering the date of the baseline interview. During these calls, the interviewers followed the same procedure mentioned above. Again, we considered a lost record if the older adult did not answer the call twice on two days. Emotional health outcomes were asked directly using the questionnaires, but in a different order than before, with items in different places. At the end of the interview, they asked about vaccination against COVID-19.

Trained interviewers registered the collected information through the Google Form platform using the identification code of each participant. The principal investigator monitored this database every two days, looking for errors during data collection. In case of suspecting a wrong registration, we coordinated with the responsible interviewer to evaluate the need to re-register said entry.

### Data analysis

All the information collected was automatically recorded in a Microsoft Excel sheet (Microsoft, WA, United States). Before the analysis, we joined the baseline database with the follow-up considering the identification code of each older adult interviewed. We reviewed the database for inconsistencies in responses about vaccination. We considered the response in the baseline measurement as the valid one if it was inconsistent with the responses during the follow-up measurement. Then, we performed the descriptive analysis of the results in the total number of recruits and separately according to the vaccination status against COVID-19. We describe the relative and absolute frequencies of the qualitative variables and the mean ± standard deviation of the quantitative variables. The baseline prevalence of altered emotional health outcomes and their 95% confidence intervals (95% CI) were plotted for the total sample and separated according to COVID-19 vaccination status. We calculated the 95% CI with the logit adjust method for the design degrees of freedom (Dean and Pagano, 2015).

We assessed, in the baseline, the association between the vaccination situation and the outcomes in emotional health. We compared medians and interquartile ranges of emotional health outcome scores with the Kruskal Walis test and the frequency of altered emotional health outcomes with the Chi-square test. We estimated the odds ratios and 95% CI for having adjusted altered emotional health outcomes (aOR) by sex, age, time in days since receiving the last dose of vaccine (unvaccinated were assigned with zero), living with someone, comorbidity, history of emotional health, and history of COVID-19, using a logistic regression model.

Finally, we assessed the association between vaccination status and altered emotional health outcomes, performing multilevel logistic regression models with mixed effects with a three-level structure. In addition to the first individual level of each measurement, the healthcare network and each individual were considered as the other two levels. This analysis allows us to calculate the effects considering the stratum of the care network and the correlations of the two responses over time within the same individual, thus allowing a longitudinal and stratified assessment. In addition, we adjusted the regression model for the confounding variables: time in days since receiving their last dose of vaccine, sex, age, living with someone, comorbidity, history of mental health disease, history of COVID-19, and time in days since the basal measurement (The basal measurement records had a value of 0).

In all the previously mentioned analyses, we consider the study’s sample weights and the stratum using the svyset command. A value of p<0.05 was considered statistically significant to reject the null hypothesis in all statistical tests. Statistical software STATA MP v17 (StataCorp, Texas, USA) was used for the analysis.

### Ethics

Each participant gave verbal informed consent before being included in the study. The anonymity of the interviewees was always maintained, assigning each one an identification code. Therefore, the survey didn’t collect personally identifiable information. In cases where a participant had an acute event in her mental health, the interviewer immediately referred the participant to a psychiatrist free of charge who managed the event by telephone. The protocol is registered in the PRISA repository of the Peruvian National Institute of Health (ID code: EI00000001999), and it was approved by the Institutional Review Board of the Instituto Nacional del Corazón - EsSalud (Certificate of approval 25/2021-CEI).

## 3. Results

### Recruitment and baseline measurement

We randomly selected and invited 1,686 older adults to the study. 51.1% (n = 861) of them met the selection criteria for baseline measurement. Among the reasons for not participating in the study were not responding to the call (n=695), having been diagnosed with COVID-19 in the last three months (n=57), refusing to participate (n=51), not wanting to be vaccinated (n=11), have symptoms related to COVID-19 at the time of the interview (n=8) and have been vaccinated in another country (n=3). Then, 20.8% (n = 179) refused to participate or did not respond to the call for follow-up measurement (Figure 2). The frequency distribution of gender and mean age were similar between those who did not participate in the baseline measurement or during the follow-up measurement compared to the recruited patients (Table S1 in Supplementary results)

**Figure 2.**
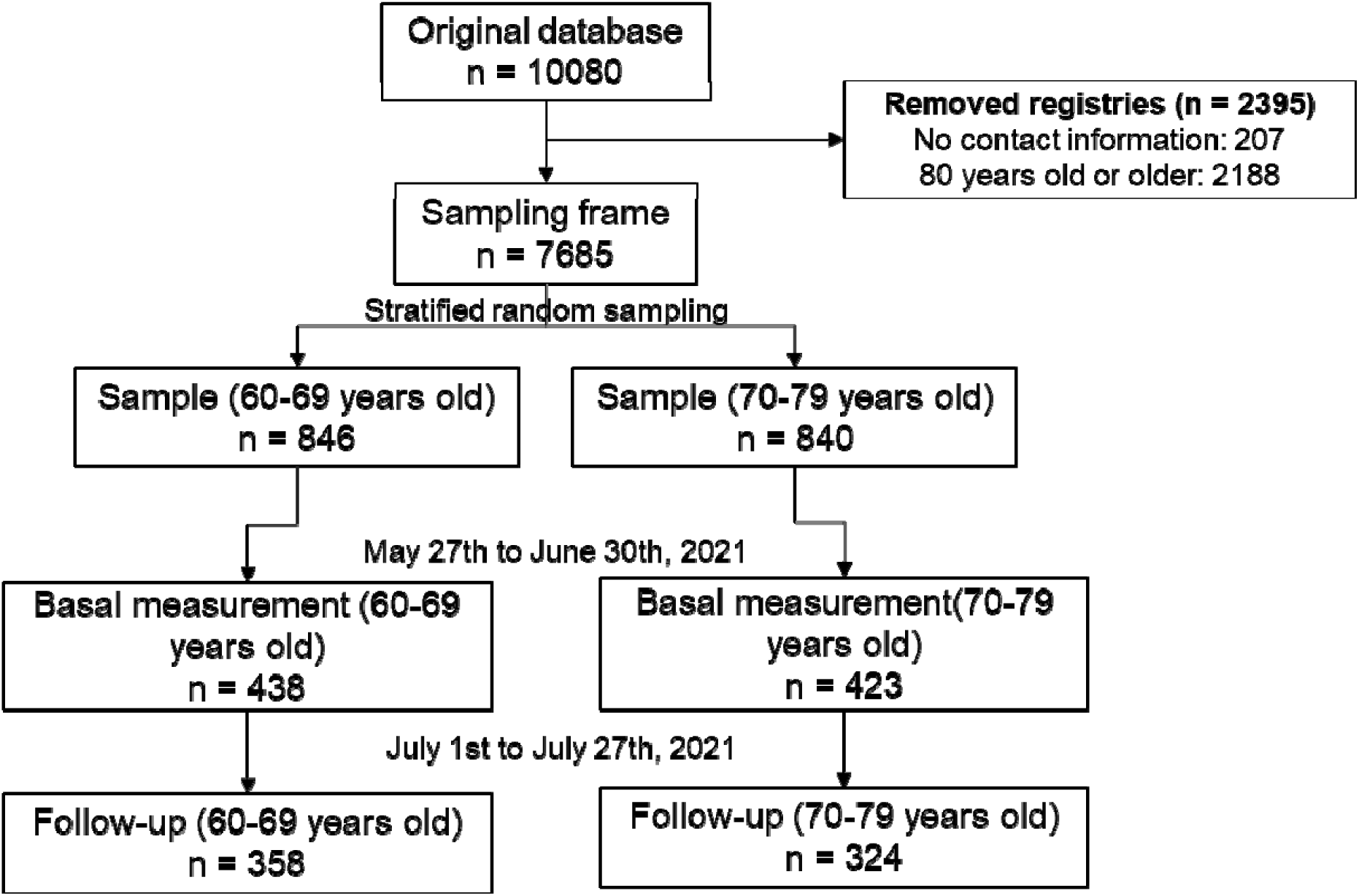
Participant’s selection flowchart.

The main characteristics of older adults aged 60 to 79 years affiliated with EsSalud’s CAMs are in Table 1. 54.5% of widowers and divorcees older adults had been in that status for less than a year. Since the pandemic’s start, 20.8% have been hospitalized among those ever diagnosed with COVID-19. On the other hand, among older adults who had at least one family member, whom they live, with a diagnosis of COVID-19, since the start of the pandemic, 17.2% of them had at least one family member who died due to COVID-19.

**Table 1.** Baseline characteristics of older adults recruited from May 27th to June 30th, 2021: Total and according to vaccination status (n=861)^†^.

| Characteristics | n | Weighted % | Unvaccinated (n=67) |  | One dose of vaccine (n=395) |  | Two doses of vaccine (n=399) |  |
| --- | --- | --- | --- | --- | --- | --- | --- | --- |
| **Age (years)<sup>‡</sup>** | 71.5 | 72.2 | 68.2 | 0.65 | 71.4 | 0.2 | 73.4 | 0.19 |
| **Women** | 655 | 75.7 | 50 | 75.6 | 308 | 77.7 | 297 | 73.9 |
| **Civil status** |  |  |  |  |  |  |  |  |
| Single | 101 | 11.4 | 6 | 8.3 | 55 | 13.7 | 40 | 9.7 |
| Married or living with partner | 507 | 58.5 | 41 | 61.8 | 223 | 54.7 | 243 | 61.4 |
| Widowed | 211 | 25.7 | 15 | 24.8 | 93 | 25.7 | 103 | 25.8 |
| Divorced | 40 | 4.5 | 4 | 5.2 | 24 | 5.9 | 12 | 3.1 |
| **Living with someone<sup>£</sup>** | 775 | 90.9 | 60 | 91.1 | 350 | 87.8 | 365 | 93.7 |
| **Geriatric Comorbidity Index<sup>£</sup>** |  |  |  |  |  |  |  |  |
| No comorbidities | 286 | 34.3 | 21 | 30.6 | 111 | 28.5 | 154 | 40.0 |
| Class I | 72 | 8.1 | 9 | 12.5 | 36 | 8.5 | 27 | 7.1 |
| Class II | 252 | 26.9 | 17 | 24.4 | 125 | 28.4 | 110 | 25.9 |
| Class III | 137 | 16.8 | 12 | 20.7 | 62 | 16.9 | 63 | 16.1 |
| Class IV | 114 | 14.0 | 8 | 11.8 | 61 | 17.7 | 45 | 10.9 |
| **Mental health disease history** | 56 | 5.4 | 7 | 9.8 | 25 | 5.5 | 24 | 4.8 |
| **Psychotherapy history<sup>\*\*</sup>** | 21 | 38.2 | 4 | 55.2 | 9 | 37.4 | 8 | 34.2 |
| **Psychotropic drug history<sup>\*\*</sup>** | 29 | 56.9 | 6 | 85.4 | 12 | 60.0 | 11 | 46.2 |
| **COVID-19 history<sup>£</sup>** | 87 | 9.6 | 10 | 13.6 | 53 | 12.9 | 24 | 6.1 |
| **COVID-19 history in family<sup>\*\*\*‡</sup>** | 123 | 14.8 | 16 | 25.5 | 69 | 19.2 | 38 | 9.5 |

<sup>†</sup>Some variables do not have 861 observations due to missing data; <sup>‡</sup>Absolute mean and weighted mean; <sup>\*\*</sup>Proportion based on total people with a mental health disease history; <sup>\*\*\*</sup>Proportion based on the total number of people living with someone; <sup>‡</sup>p<0.001; <sup>£</sup>p<0.05

Regarding the vaccination situation, in the baseline measurement, 43.9% of the respondents received only one dose with an average time of 15.6 days from the date they received the vaccine, and 49.1% received the two doses with an average time of 16.9 days from the date you received the vaccine. On the other hand, 5.4% of older adults reported having a previous diagnosis of a mental health disorder. However, we found the prevalence of general anxiety, assessed with GAD-2, to be 16.4% (95% CI: 14.1 to 19.1), and the prevalence of general depression, estimated by PHQ-2, was 8.0% (95% CI: 6.3 to 10.0). In addition, we found that those who had two doses of the vaccine had less fear of COVID-19, anxiety for COVID-19, worry for COVID-19, and general anxiety compared to those who were unvaccinated or those who had a vaccine. However, we didn’t observe a trend in the case of the outcome of general depression (Figure 1).

**Figure 2.**
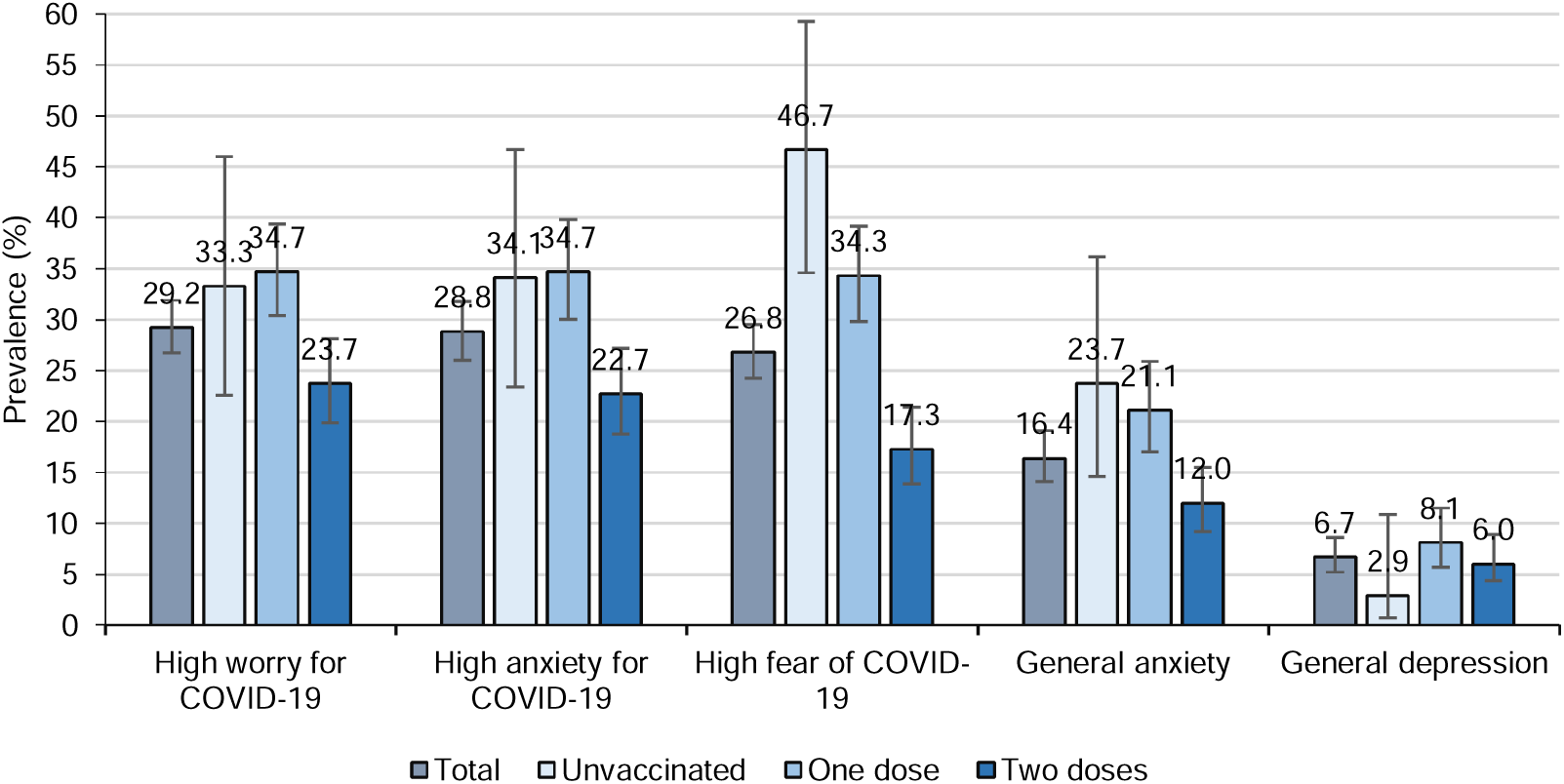
Baseline prevalence of emotional health outcomes in total and according to vaccination status in older adults recruited from May 27th to June 30th, 2021 (n=861)

### Association between vaccination status and emotional health outcomes at baseline

During the baseline measurement, we observed that older adults who had two doses of the vaccine had less fear of COVID-19 (aOR: 0.27; 95% CI: 0.13 to 0.55) than those who were unvaccinated. Similarly, we observed a lower anxiety for COVID-19, worry for COVID-19, and general anxiety in those who had two doses of the vaccine, but without statistical significance. Meanwhile, those who had one or two doses of the vaccine were more likely to have general depression than those who were unvaccinated, although this was not statistically significant (Table 2).

**Table 2.** Association between vaccination status and emotional health outcomes during baseline measurement (n=861)

| Outcomes | Vaccination against COVID-19 status |  |  |
| --- | --- | --- | --- |
|  | Unvaccinated (n=67) | One dose (n=395) | Two doses (n=399) |
| **Fear of COVID-19 (FCV-19S)** |  |  |  |
| Median and IQR score | 10.5 (7 a 17) | 8 (7 a 15) | 7 (7 a 10) |
| aOR (95% CI) for high fear of COVID-19 (twelve points or more) | Ref. | 0.65 (0.34 a 1.27) | 0.27 (0.13 a 0.55) |
| **Anxiety for COVID-19 (CAS)** |  |  |  |
| Median and IQR score | 0 (0 a 2) | 0 (0 a 1) | 0 (0 a 0) |
| aOR (95% CI) for high anxiety for COVID-19 (one point or more) | Ref. | 1.18 (0.61 a 2.26) | 0.67 (0.34 a 1.30) |
| **Worry for COVID-19 (PRE-COVID-19)** |  |  |  |
| Median and IQR score | 6 (8 a 15.5) | 6 (8 a 14) | 6 (6 a 11) |
| aOR (95% CI) for high worry for COVID-19 (twelve points or more) | Ref. | 1.21 (0.60 a 2.44) | 0.74 (0.36 a 1.52) |
| **General anxiety (GAD-2)** |  |  |  |
| Median and IQR score | 0 (0 a 1) | 0 (0 a 1) | 0 (0 a 0) |
| aOR (95% CI) for general anxiety (two points or more) | Ref. | 0.76 (0.37 a 1.57) | 0.46 (0.21 a 1.03) |
| **General depression (PHQ-2)** |  |  |  |
| Median and IQR score | 0.0 (0 a 0) | 0 (0 a 0) | 0 (0 a 0) |
| aOR (CI 95%) for general depression (three points or more) | Ref. | 2.65 (0.56 a 12.64) | 1.98 (0.40 a 9.83) |

IQR: Interquartile range; aOR: Adjusted odds ratio for days since receiving the last vaccine dose, sex, age, living with someone, comorbidity, history of mental health disease, and history of COVID-19 diagnosis; 95% CI: 95% confidence interval; CAS: Coronavirus Anxiety Scale; FCV-19S: Fear of COVID-19 Scale; GAD-2: Generalized Anxiety Disorder; PHQ-2: Patient Health Questionnaire; PRE-COVID-19: Scale to measure worry for contagion of the COVID-19.

### Association between vaccination and mental health problems at one month of follow-up

The mean follow-up time for 661 older adults was 31.4 ± 0.14 days. Considering the one-month follow-up period, we observed that older adults with two doses of the COVID-19 vaccine had less fear of COVID-19 (ORa: 0.19; 95% CI: 0.07 to 0.51) and less anxiety for COVID-19 (aOR: 0.45; 95% CI: 0.22 to 0.89), compared to those who were unvaccinated. We observed similar results in the outcomes of worry for COVID-19 and general anxiety; however, there is high uncertainty about these estimates (Table 3).

**Table 3.** Association between vaccination status and emotional health outcomes for one month follow-up (n=661)

| Outcomes | One dose of vaccine against COVID-19 |  | Two doses of vaccine against COVID-19 |  |
| --- | --- | --- | --- | --- |
|  | aOR | 95% CI | aOR | 95% CI |
| **Fear of COVID-19 (FCV-19S)** |  |  |  |  |
| High fear (twelve points or more) | 0.55 | (0.22 a 1.39) | 0.19 | (0.07 a 0.51) |
| **Anxiety for COVID-19 (CAS)** |  |  |  |  |
| High anxiety for COVID-19 (one point or more) | 0.94 | (0.49 a 1.80) | 0.45 | (0.22 a 0.89) |
| **Worry for COVID-19 (PRE-COVID-19)** |  |  |  |  |
| High worry for COVID-19 (twelve points or more) | 1.26 | (0.52 a 3.06) | 0.72 | (0.29 a 1.81) |
| **General anxiety (GAD-2)** |  |  |  |  |
| General anxiety (two points or more) | 0.84 | (0.42 a 1.69) | 0.51 | (0.24 a 1.07) |
| **General depression (PHQ-2)** |  |  |  |  |
| General anxiety (three points or more) | 1.69 | (0.32 a 9.00) | 1.16 | (0.23 a 5.97) |

Reference category: Unvaccinated; aOR: Adjusted odds ratio for days since receiving the last vaccine dose, sex, age, living with someone, comorbidity, history of mental health disease, and history of COVID-19 diagnosis and follow-up time in days; 95% CI: 95% confidence interval; CAS: Coronavirus Anxiety Scale; FCV-19S: Fear of COVID-19 Scale; GAD-2: Generalized Anxiety Disorder; PHQ-2: Patient Health Questionnaire; PRE-COVID-19: Scale to measure worry for contagion of the COVID-19.

## 4. Discussion

### Summary of results

We hypothesize that vaccination has a causal effect in reducing fear, anxiety, and worry for COVID-19 and, in general anxiety and depression. Our study, conducted in a cohort from a nationally representative sample of older adults affiliated to EsSalud, partially confirmed our hypothesis. We found evidence that receiving two doses of the COVID-19 vaccine, compared with unvaccinated, reduced the likelihood of high levels of fear and anxiety for COVID-19. In contrast, we were unable to confirm these findings for any outcome in those who had received only one dose of the vaccine. To our knowledge, this is the first study to assess the effect of vaccination on emotional health in a representative sample of older adults, using novel COVID-19 perception outcomes.

### Effect of vaccination against COVID-19 in the emotional health of older adults

High fear and anxiety for COVID-19 significantly decreased in those who had two doses of the COVID-19 vaccine than those who were unvaccinated. Interestingly, the scales that measured both constructs (FCV-19S and CAS, respectively) focus mainly on the emotional and physical reaction to thoughts related to COVID-19 and its possible contagion (Lee, 2020; Mertens et al., 2021). However, the PRE-COVID-19 scale, which measured the worry for COVID-19, focused on daily dysfunction caused by thoughts about the possibility of getting COVID-19 (Caycho-Rodríguez et al., 2021c). This difference is relevant, as it would mean that vaccination could affect older adults to improve their mental well-being by reducing more intense psychosomatic symptoms of stress related to the pandemic (fear/anxiety of COVID-19) (Chandu et al., 2020); but without reducing daily thoughts and behaviors associated with the possibility of contagion (COVID-19 concern).

On the other hand, although we observed a slight decrease in general anxiety, the effect of vaccination on general depression and anxiety outcomes, measured with PHQ-2 and GAD-2, respectively, was not significant. However, previous studies conducted in adults from the United States (Perez-Arce et al., 2021), Turkey (Bilge et al., 2021), and China (Yuan et al., 2021), and health professionals from Turkey (Sugihara et al., 2021) reported that those with at least one dose of the vaccine against COVID-19 have lower scores on the depression scales, measured with the PHQ-4, PHQ-9, and the Beck Depression Inventory, and on anxiety scales, measured with the GAD-7 and the Beck Anxiety Inventory. The mechanisms and causes of depression and anxiety in older adults are related to psychosocial factors of loneliness and lost, and neuroendocrine and vascular disorders (Alexopoulos, 2019; Hellwig and Domschke, 2019). The previously mentioned studies included adults in general with low representation of older adults, so, according to our results, the effect of vaccination would not be sufficient to significantly reduce these outcomes in emotional health in older adults, since they would respond to other factors intrinsic and extrinsic that were not measured in the present study.

On the other hand, the evaluation of the effect of vaccination against COVID-19 on emotional health could be affected by the perception of the vaccine’s effectiveness. Older adults are particularly susceptible to fake news or misinformation (Moore and Hancock, 2020), which could influence their perception of vaccination against COVID-19 and diminish its effect on their mental health. This effect should be evaluated in future studies. Similarly, the perception of effectiveness and vaccination intention could also be affected by the type of vaccine manufacturer, with the BNT162b2 vaccine (BioNTech, Pfizer) being the most preferred, and the ChAdOx1-S vaccine (Oxford, AstraZeneca) the least preferred in developed countries (Merkley and Loewen, 2021; Quito, 2021; YouGov, 2021). Thus, even though most older adults were vaccinated with the BNT162b2 vaccine (BioNTech, Pfizer), the political scandal in Peru regarding the BBIBP-CorV (Sinopharm) vaccine (Mayta-Tristán and Aparco, 2021) may have partially affected the effect of vaccination on the emotional health of this population.

### Second dose of vaccine against COVID-19

We observe the effect on emotional health from the second dose and not in those who have only one dose of the vaccine. This result is different from previous studies where all those who had at least one dose were included in the vaccinated group, regardless of whether they had both doses or not (Bilge et al., 2021; Perez-Arce et al., 2021; Sugihara et al., 2021; Yuan et al., 2021). Clinical effectiveness studies have shown the need for a second dose of the vaccine to have greater effectiveness in preventing mortality and severe disease from COVID-19 in older adults (Bernal et al., 2021; Kissling et al., 2021). This information was communicated promptly to the population, making most people aware of the need for a second dose, especially those willing to be vaccinated (Goldfarb et al., 2021; Stead et al., 2021). This may explain that in the context where the communicational emphasis was placed on the need for the second dose of the COVID-19 vaccine, the effect on emotional health could mainly be observed in those who received the two doses of the vaccine. Considering the high proportion of older adults with two doses of the vaccine (Kriss et al., 2021), the effect of vaccination reported in other studies may have been carried by those who had both doses, compared to those who had only one dose of the vaccine against COVID-19.

However, the presence of new variants of concern, such as B.1.1.529, could affect the population’s mental health (Jain and Jolly, 2021; Su et al., 2022) due to their greater infectivity and immune escape from vaccination (Ren et al., 2022). Given this, the need for a third (Eroglu et al., 2022; World Health Organization, 2021), or even a fourth (Tylicki et al., 2022), COVID-19 vaccine booster is currently under discussion. So, considering that the perception of the vaccine’s effectiveness correlates with the level of concern about the new variants (Temsah et al., 2021), it is important to continue monitoring mental health in the most vulnerable populations such as the elderly, and its evolution during future vaccination policies against COVID-19.

### Public health relevance

As of January 2022, the two-dose vaccination rate in adults aged 60 years and older in different countries was around 80% (Ritchie et al., 2020). Among the reasons for older adults to decide to be vaccinated is the fear of developing the disease and the perception of the vaccine’s effectiveness to prevent the disease (Bhanu et al., 2021; Caycho-Rodríguez et al., 2021a). However, the lack of reliable information, the fear of possible adverse effects, and the limited access to receive the vaccines mean that many older adults do not get vaccinated or do not have the opportunity to get vaccinated (Bhanu et al., 2021). In this sense, the communication strategy to promote vaccination against COVID-19 could be complemented with the message of reducing fear and anxiety about being infected with COVID-19. Thus, integrating with other elements necessary to have an adequate vaccination rate, such as the empowerment of the first level of care and the availability and access to vaccines (Castillo et al., 2021), could improve citizen confidence in the vaccination process.

Currently, the COVID-19 pandemic has a significant impact on mental health, which will continue in the medium and long term (Esterwood and Saeed, 2020). From the point of view of positive epidemiology, our results propose vaccination against COVID-19 as a positive determinant of mental health in older adults (VanderWeele et al., 2020). So, vaccination against COVID-19 could contribute to the partial improvement of the emotional health of older adults. However, we must consider that the mental health of older adults depends on various intrinsic and extrinsic factors that not only respond to the COVID-19 pandemic (Alexopoulos, 2019; Hellwig and Domschke, 2019).

### Limitations and strengths

The interpretation of the results of this study must consider the following limitations. First, we were unable to obtain the planned sample size for the primary analysis, making the statistical power of our results insufficient to find statistically significant results. Thus, we do not rule out the effect of the vaccine on worry for COVID-19 and general anxiety, which should be evaluated in future studies. In addition, the low representativeness of the older adults affiliated with EsSalud registered in the CAM database, the high refusal to participate in the study, and the loss during follow-up could have caused selection bias. Thus, it is likely that older adults in CAMs have greater access to receiving the vaccine and to activities that improve their mental health, so the effect that we measured in the study may be overestimated. Additionally, the way to determine the vaccination status was by self-report, so the measurement of this variable could have been overestimated due to the social desirability bias. On the other hand, even though we did not conduct clinical interviews to evaluate the emotional health in the present study, we used different specific psychometric instruments for the perception of COVID-19 and general anxiety and depression. These tools have robust evidence of psychometric validity in our population of interest, making reliable the constructs that we measured.

## Conclusion

Vaccination against COVID-19 with two doses in older adults reduces fear and anxiety for COVID-19, compared to those who were unvaccinated. However, we observed no effect in general anxiety and general depression, nor in those who only had one vaccine dose.

## Data Availability

All data produced in the present study are available upon reasonable request to the authors

## Abbreviations

CAM: Centers for the eldery
CAS: Coronavirus Anxiety Scale
EsSalud: Social Health Insurance of Peru
FCV-19S: Fear of COVID-19 Scale
GAD-2: Generalized Anxiety Disorder
PHQ-2: Patient Health Questionnaire
PRE-COVID-19: Scale to measure worry for contagion of the COVID-19

## Acknowledgments

To the workers from Gerencia de la Persona Adulta Mayor y Prestaciones Sociales, EsSalud for participating in the data collection process.

